# Progesterone/Estradiol Ratio Is Related to Real-Life Alcohol Consumption in Alcohol Use Disorder in a Sex- and Menstrual Cycle Phase-Dependent Manner

**DOI:** 10.1101/2022.12.21.22282762

**Authors:** Sabine Hoffmann, Sarah Gerhardt, Christiane Mühle, Iris Reinhard, Dominic Reichert, Patrick Bach, Rafat Boroumand-Jazi, Christine Kuehner, Alvaro Aguilera, Acelya Aslan, Nadja S. Bahr, Matthew Belanger, Friederike Deeken, Claudia Ebrahimi, Pascale C. Fischbach, Marvin Ganz, Maria Garbusow, Charlotte M. Großkopf, Marie Heigert, Angela Hentschel, Damian Karl, Shuyan Liu, Massimiliano Mazza, Patricia Pelz, Mathieu Pinger, Matthias Reichl, Carlotta Riemerschmid, Annika Rosenthal, Johannes Steffen, Jens Strehle, Friederike Wedemeyer, Franziska Weiss, Julia Wenzel, Gesine Wieder, Alfred Wieland, Judith Zaiser, Hilmar Zech, Sina Zimmermann, Johannes Kornhuber, Christian P. Müller, Wolfgang H. Sommer, Rainer Spanagel, Tobias Banaschewski, Lorenz Deserno, Ulrich W. Ebner-Priemer, Herta Flor, Peter Kirsch, Marcella Rietschel, Sabine Vollstädt-Klein, Henrik Walter, Andreas Meyer-Lindenberg, Michael A. Rapp, Stephanie Witt, Michael N. Smolka, Andreas Heinz, Heike Tost, Falk Kiefer, Markus Reichert, Bernd Lenz, the ReCoDe-Consortium

**Author notes:** Corresponding author at: Department of Addictive Behavior and Addiction Medicine, Central Institute of Mental Health (CIMH), Medical Faculty Mannheim, Heidelberg University, J 5, 68159 Mannheim, Germany. Markus Reichert and Bernd Lenz have contributed equally to this work.

## Abstract

**Background:** Alcohol use disorder (AUD) is a critical public health issue with sex-specific characteristics and the need for a better mechanistic understanding. Initial evidence suggests that progesterone can reduce alcohol intake, while estradiol leads to an increase. However, we lack knowledge about how progesterone relative to estradiol influences alcohol use patterns in females and males with AUD.

**Methods:** This multicenter within-subject study analyzed data on real-life alcohol use (21,438 intensively-sampled smartphone entries), menstrual cycle, and serum progesterone/estradiol ratios (677 blood samples) gathered during a 12-month follow-up in 74 naturally cycling females and 285 males with AUD (mean age: 29.7 and 37.8 years, respectively; data collection: 2020–2022). We used multilevel modelling to identify changes in alcohol use and progesterone/estradiol ratios across the menstrual cycle in females and associations between progesterone/estradiol ratios and alcohol use in males.

**Results:** During the late luteal phase, females showed 0.6- to 0.8-fold lower (predicted) probabilities of binge drinking and 2.8- to 5.6-fold higher mean progesterone/estradiol ratios compared to the menstrual, follicular, and ovulatory phases. Similarly, in males, an increase of 10 units in the progesterone/estradiol ratio was related to 8 and 9% lower probabilities of binge drinking and any alcohol use, respectively.

**Conclusions:** Based on ecologically valid results, this study reveals that higher progesterone/estradiol ratios can protect against problematic alcohol use in females and males with AUD. Therefore, the progesterone/estradiol ratio is a promising treatment target. Translated into clinical practice, our results also indicate that females with AUD may benefit from menstrual cycle phase-tailored treatments.

## Introduction

Alcohol use disorder (AUD) and excessive alcohol use are serious public health threats and impose a high burden on individuals, their families, and society as a whole (1, 2). The prevalence rates of AUD are estimated to be between 4% and 5% in females^1^ and between 12% and 15% in males in the Americas and Europe (2). Although AUD is more prevalent in males, females progress more rapidly to AUD from the onset of alcohol drinking (3). To develop more effective treatment strategies, a better understanding of the mechanisms underlying AUD is crucial.

The sex differences in problematic alcohol use (2-4) and the effects of sex hormones on the brain reward system as established in animal experiments (5-8) indicate that progesterone and estradiol might play a key role in AUD of females and males. A systematic review (9) reports that higher estrogen levels are related to increased alcohol use in females with inconsistent results in males. Moreover, this review concludes that so far minimal is known about how progesterone is associated with alcohol use in humans. Very recently, we found a correlation between high progesterone serum levels and lower craving for alcohol in females with AUD (10).

For cocaine use disorder, results from animal studies suggest that in both sexes the administration of progesterone may serve as a potential treatment, whereas estradiol leads to an increase in the risk of addictive behaviors (11, 12). In female and male rodents, progesterone attenuates the reinforcing effects of cocaine with reduced acquisition, escalation, and reinstatement of self-administration (13). In cocaine-dependent human females, high progesterone levels are associated with low stress- and cue-induced craving and anxiety (14), and the administration of progesterone to cocaine-dependent females and males leads to an increase of its metabolite, allopregnanolone, in plasma with a reduction in craving, normalization of stress reactivity, and improvement of mood and cognitive performance (15, 16). Interestingly, estradiol sensitizes dopamine neurons in the ventral tegmental area (17) and increases cocaine-induced dopamine levels in the dorsal striatum of female rats (18), two key areas involved in the neurocircuitry of addictive behavior. Indeed, due to its therapeutic potential, progesterone is of particular interest. However, it remains unclear whether the indicated protective effects of progesterone related to cocaine use are also valid in individuals with AUD.

The menstrual cycle has been used as a naturalistic setting to investigate behaviors related to the varying brain exposures to progesterone and estradiol in female samples (19-21) (low progesterone and estradiol during the follicular phase, low progesterone and peaking estradiol during the ovulatory phase, peaking progesterone and mildly elevated estradiol during the luteal phase, and falling progesterone and estradiol before the menstrual phase). In a tightly controlled within-person study of 22 naturally cycling young female university students, Martel et al. (22) found a lower likelihood of alcohol drinking in phases of high progesterone relative to estradiol saliva levels (during the luteal phase). These effects were amplified for binge drinking (versus any alcohol use) and present during weekend days (but not during weekdays). In line with the above reported results, these observations suggest that progesterone may cause a decrease in problematic alcohol use, while estradiol causes an increase. However, two systematic reviews concerning changes in alcohol use across the menstrual cycle concluded that the data are inconsistent and subject to several limitations (23, 24). First, most of the research used convenience samples of college students with narrow age ranges, investigated small samples, and was published before the year 2000. Second, a comparison of the studies is limited due to inconsistent methods adopted to operationalize menstrual cycle phases, unstandardized cycle phase designations (in length and position throughout the menstrual cycle), and varying definitions of alcohol use (often retrospective self-reports that are subject to recall biases (25)). Moreover, some studies did not control for use of hormonal contraceptives, which more than 550 million females used worldwide by 2019 (26).

Overall, the existing literature on the effects of progesterone and estradiol on alcohol use patterns is limited. In particular, there is a research gap addressing associations between activities of progesterone relative to estradiol (namely, the progesterone/estradiol ratio) and alcohol use patterns in females and males with AUD. This gap appears highly critical against the background that previous research highlighted the general importance of the progesterone/estradiol ratio for studying behavior and physiology (27-30). Similarly, we lack conclusive evidence on how alcohol use varies across the menstrual cycle in females with AUD and whether the progesterone/estradiol ratio is also related to alcohol use patterns in males with AUD (which is very likely since progesterone and estradiol are also relevant to the male organism (31-33)).

### Aims of the Study

Although sex disparities in alcohol use have decreased during the last decades (34), females are still highly underrepresented in AUD research. Thus, separate investigations for females and males are necessary to avoid sex bias (35, 36). In this study, we investigated activities of the progesterone/estradiol ratio in relation to alcohol use in both sexes and tested the following hypotheses: (1) In premenopausal naturally cycling females with AUD, we used the menstrual cycle as a proxy for longitudinal changes in progesterone/estradiol ratios. We expected a lower probability of binge drinking because of higher mean progesterone/estradiol ratios during the luteal phase than during the ovulatory phase. (2) In males with AUD, we hypothesized that higher progesterone/estradiol ratios are associated with lower likelihoods of binge drinking and any alcohol use.

Previous research indicates that the effects of progesterone and estradiol activities on alcohol use patterns differ between weekend days and weekdays (22) with weekend status as a permissive factor in the association between hormones and alcohol drinking and between varying severities of AUD with stronger effects in mild AUD (37). Therefore, we investigated a sample of females and males with prevalently mild-to-moderate AUD and explored whether the interactions of menstrual cycle phases and the progesterone/estradiol ratio with weekend days versus weekdays and AUD severity affect alcohol use patterns.

## Methods and Materials

### Study Design and Sample Description

This 12-month follow-up cohort study was conducted within the frame of the Collaborative Research Centre Transregio 265 (TRR265) (38). Between February 2020 and July 2022, we screened 3,492 individuals, and of these, 672 females and males with mainly mild-to-moderate AUD were enrolled at three study sites in Germany (Charité – Universitätsmedizin Berlin, Technische Universität Dresden, and Central Institute of Mental Health (CIMH) Mannheim; TRR265 cohort; preregistration: DRKS00020580). The study design included four visits (baseline and follow-ups scheduled at 4, 8, and 12 months after enrollment) and real-life Ecological Momentary Assessments (EMA) every other day (39). The project was approved by the local ethics committees of Charité – Universitätsmedizin Berlin, Technische Universität Dresden, and Heidelberg University.

Participants were included if they fulfilled the criteria of AUD (that is ≥2) according to the Diagnostic and Statistical Manual of Mental Disorders, Fifth Edition (DSM-5) (40), were between 16 and 65 years old, were able to understand the study protocol, and gave written informed consent. Exclusion criteria comprised the intake of central nervous system agents in the last 10 days or less than 5 half-life periods ago, contraindications for magnetic resonance tomographic scans, DSM-5 bipolar disorder, psychotic disorder, schizophrenia or disorder from the schizophrenic spectrum, or substance addiction except for alcohol, nicotine, and cannabis in medical history, severe head injury or other severe diseases of the central nervous system in medical history, pregnancy, and breastfeeding.

Seventy-four females were classified as premenopausal naturally cycling females. They reported a regular cycle during the previous 3 months of at least 21 and at maximum 35 days, negated intake of contraceptives or other hormones, reported the date of the last menses, and were not older than 42 years. This maximum age results from the age of 50 years (= median age at cessation of menses in females in Germany (41, 42)) minus (up to) 8 years for perimenopause (43). We also included 285 males.

### Data Sampling: Ecological Momentary Assessment

Participants were asked to submit information through a smartphone e-diary application every second day (for further details see (39)). They were reminded by an alarm at noon, which could be postponed until 8 PM. For this project, we analyzed the e-diary entries on alcohol use: *Think about yesterday. Which and how many alcoholic drinks did you consume? Think about the day before yesterday. Which and how many alcoholic drinks did you consume?* The participants selected alcoholic drinks from a list with information on volume and alcohol content. In the sample of premenopausal naturally cycling females with AUD, we analyzed 4,887 entries of 115 observation periods from 74 females (baseline: 69; 4-month follow-up: 26; 8-month follow-up: 12; 12-month follow-up: 8), which were coupled with two complete menstrual cycles oriented to the onset of the last menses. Hence, we analyzed two menstrual cycles after the baseline assessment, one menstrual cycle before and one after the 4-month and 8-month assessments, and two menstrual cycles before the 12-month assessment. Similarly, in the male sample, we used 2-month entries distributed around the four study visits (N = 16,597) as shown in Supplementary Figure S1. At the median, the participants answered 75% of the queries on their alcohol use (interquartile range 46 – 90) indicating sound compliance rates given the long-term assessment in real-life.

### Classification of Menstrual Cycle Phases

At every study visit, females were asked to answer the following questions (44, 45): *During the previous three months, did you have a regular cycle? Yes/No. How long does your cycle last? Less than 21 days/21 to 23 days/24 to 26 days/27 to 29 days/30 to 32 days/33 to 35 days/more than 35 days. When did your last menstruation start? Day*|*Month*|*Year. Do you take contraceptives or other hormones? Yes/No*. To classify the menstrual cycle phase, we used a counting approach recently recommended (46). The method takes individual differences in cycle length into account, which are mostly due to variances in the length of the follicular phase (47). For further details, see Supplementary Table S1.

### Quantification of Progesterone and Estradiol Serum Concentrations

Blood was drawn during every study visit. After incubation for 0.5 to 2 h at room temperature, the vials were centrifuged for 15 min at 1,100 g and 4°C. Then, serum aliquots were transferred to −80°C. All samples were jointly stored at the Biobank of Psychiatric Diseases Mannheim (48). We used the following competitive enzyme-linked immunosorbent assays (ELISAs): (1) Progesterone ELISA (RE52231; IBL International GmbH, Hamburg, Germany) and (2) Estradiol ELISA (EIA-2693; DRG Instruments GmbH, Marburg, Germany). In parallel to standard curves on every 96-well plate ranging from 0.15 to 40 ng/mL for progesterone and from 12.5 to 2000 pg/mL for estradiol, we dispensed duplicates of 25 µL of serum for the progesterone ELISA and 25 µL of serum for the estradiol ELISA. The intra- and inter-assay coefficients of variation were 3.0% and 10.2% for the progesterone ELISA and 7.0% and 12.9% for the estradiol ELISA, respectively. For statistical analyses, we used the progesterone (ng/mL)*1000/estradiol (pg/mL) ratios of 677 blood samples (mean ± standard deviation and range; 72 samples from 49 females: 45.0 ± 61.4, 1.5 – 334.4; 605 samples from 285 males: 12.8 ± 8.5, 1.5 – 114.4).

### Statistical Analysis

We estimated multilevel (mixed) models, including random intercepts, to account for the nesting structure of each data point within a participant. Generalized linear mixed models were fitted for binge drinking and any alcohol use on a daily basis as binary dependent variables (yes versus no) with menstrual cycle phase (or cycle day) in premenstrual naturally cycling females or progesterone/estradiol ratios in males as predictors (main effects). A day was classified as a binge drinking day when the females or males reported at least 4 or 5 standard drinks, respectively (4). A standard drink was defined as 12 g of pure alcohol, which refers to approximately 330 mL beer or 110 mL wine (corresponding alcohol percentages: 5% and 11%). Afterwards, we tested for differential effects of the menstrual cycle phase (or cycle day) or progesterone/estradiol ratios by incorporating interaction effects with AUD criteria and weekend days versus weekdays in the models. In this study, we classified Friday to Sunday as weekend days and Monday to Thursday as weekdays. To compute the cycle day with minimum binge drinking risk, we estimated a model with squared menstrual cycle day added as a predictor (instead of cycle day or cycle phases alone). Moreover, we used general linear mixed models to evaluate the effects of menstrual cycle phases on the progesterone/estradiol ratios as an outcome variable. The number of fulfilled AUD criteria and weekend days versus weekdays were included in these models, since these parameters are associated with alcohol use in our sample (39). To account for hormonal variations over the day, we also included the time of blood collection in the mixed models. Data were analyzed and visualized with SPSS (IBM SPSS for Windows 27.0; SPSS Inc., Chicago, IL, USA), SAS (Version 9.4, SAS Institute Inc. Cary, NC, USA), and Graph Pad Prism 5 (Graph Pad Software Inc., San Diego, CA, USA).

## Results

### Sample Characteristics

For this project, 359 study participants (127 from Berlin, 69 from Dresden, and 163 from Mannheim) were allocated to the AUD groups of premenopausal naturally cycling females and males (for sample characteristics see Table 1).

**Table 1.** Sample Characteristics

| | Premenopausal Naturally<br>Cycling Females with AUD<br>N, M ± SD, or % | Males with AUD<br>N, M ± SD, or % |
| --- | --- | --- |
| *Sample Characteristics* |  |  |
| Age (years) | 29.7 (6.9) | 37.8 (12.8) |
| Marital status |  |  |
| % Single | 67.6% | 48.0% |
| % Living in marriage or partnership | 25.7% | 42.5% |
| % Living separately | 2.7% | 4.0% |
| % Divorced | 2.7% | 5.1% |
| % Widowed | 1.4% | 0.4% |
| % Employed last three months | 87% | 81% |
| Highest school qualification |  |  |
| % Pupil at a general education school/in training | 4.1% | 3.3% |
| % Secondary general school certificate<sup>1</sup> | 0.0% | 1.8% |
| % General Certificate of Secondary Education<sup>2,3</sup> | 13.5% | 16.8% |
| % Advanced technical college certificate<sup>4</sup> | 12.2% | 9.2% |
| % General Certificate of Education<sup>5</sup> | 68.9% | 67.4% |
| % Another school degree | 1.4% | 1.5% |
| Number of fulfilled AUD criteria | 4.3 (1.6) | 4.0 (1.6) |
| % mild AUD (2 – 3 criteria met) | 37% | 44% |
| % moderate AUD (4 – 5 criteria met) | 39% | 37% |
| % severe AUD ($\geq 6$ criteria met) | 24% | 19% |
| % 3-month prevalence tobacco smoking | 11% | 7% |
| % 3-month prevalence cannabis use | 10% | 6% |
| *Real-life e-diary entries (EMA)* |  |  |
| N(cases) | 74 | 285 |
| N(observations) | 4,841 | 16,597 |
| % days binge drinking | 21% | 29% |
| % weekend days | 27% | 37% |
| % weekdays | 16% | 24% |
| % days any alcohol | 48% | 58% |
| % weekend days | 56% | 66% |
| % weekdays | 43% | 53% |
| Daily alcohol use (g/d) | 24.8 (16.7) | 39.6 (27.2) |
| Weekend days (g/d) | 31.3 (41.8) | 50.1 (56.6) |
| Weekdays (g/d) | 18.7 (30.3) | 32.4 (44.3) |
| *Blood samples and hormone measures* |  |  |
| N(cases) | 49 | 285 |
| N(observations) | 72 | 605 |
| Progesterone/estradiol ratio | 45.0 (61.4) | 12.8 (8.5) |

The table reports absolute frequencies, mean values (M), standard deviations (SD), and relative frequencies. AUD, alcohol use disorder; EMA, ecological momentary assessment; Progesterone/estradiol ratio, progesterone (ng/mL)\*1000/estradiol (pg/mL) ratio. <sup>1</sup>Corresponds to “Hauptschulabschluss”, <sup>2</sup>to “Realschulabschluss”, <sup>3</sup>to “Abschluss polytechnische Oberschule”, <sup>4</sup>to “Fachhochschulreife”, and <sup>5</sup>to “Abitur” in the German educational system.

### Premenopausal Naturally Cycling Females with AUD: Lower Binge Drinking Risk and Higher Progesterone/Estradiol Ratios in the Late Luteal than the Other Menstrual Cycle Phases

The probability of binge drinking and the mean progesterone/estradiol ratios changed significantly across the menstrual cycle phases in the group of premenopausal naturally cycling females (Table 2). Binge drinking probability was 0.6- to 0.8-fold lower during the late luteal phase (14%) than during the menstrual, follicular, ovulatory, and mid-luteal phases (18% to 22%; Figure 1). As hypothesized, the changes in binge drinking probability across the menstrual cycle phases were paralleled by alterations in the mean progesterone/estradiol ratios with a 2.8- to 5.6-fold higher value of 95 during the late luteal phase than during the menstrual, follicular, and ovulatory phases (17 to 34) and higher values during the mid-luteal than during the ovulatory and follicular phases (Supplementary Figure S2). To compute the cycle day with minimum binge drinking risk, we estimated a quadratic association and identified the minimum at 2.25 days before the onset of menses (f(x) = 0.002x^2^ + 0.009x - 1.601; Supplementary Table S2 and Supplementary Figure S3). The likelihood of days with any alcohol use did not significantly differ between the menstrual cycle phases. Moreover, alcohol use was higher during the weekend days than during weekdays (Table 2). The interactions of cycle phases with AUD criteria or weekend days versus weekdays did not reveal significant effects (data not shown).

**Table 2.** Multilevel Modeling Results in Premenopausal Naturally Cycling Females with AUD: Association of Menstrual Cycle Phases with Probability of Binge Drinking Days, Days with Any Alcohol Use, and Progesterone/Estradiol Ratios Estimated in Separate Models

| | $\beta$ [95% CI] | p | F (df1, df2) | p | OR |
| --- | --- | --- | --- | --- | --- |
| **Binge Drinking Day (yes/no)** |  |  |  |  |  |
| **N = 4,841** |  |  |  |  |  |
| Intercept | -2.66 [-3.40; -1.91] |  | 20.13 (6, 1079) | **< .001** | 0.07 |
| AUD criteria | 0.11 [-0.05; 0.26] |  | 1.87 (1, 68) | .176 | 1.11 |
| Weekend days versus weekdays |  |  | 102.15 (1, 4834) | **< .001** | 2.18 |
| Weekend days | 0.78 [0.63; 0.93] |  |  |  |  |
| Weekdays | [Reference] |  |  |  |  |
| Cycle phases |  |  | 4.38 (4, 4834) | **.002** |  |
| Menstrual | 0.30 [0.03; 0.56] | **.028** |  |  | 1.34 |
| Follicular | 0.42 [0.17; 0.67] | **.001** |  |  | 1.53 |
| Ovulatory | 0.54 [0.27; 0.81] | **< .001** |  |  | 1.72 |
| Mid-luteal | 0.33 [0.09; 0.57] | **.008** |  |  | 1.39 |
| Late luteal | [Reference] |  |  |  |  |
| **Day with Any Alcohol Use (yes/no)** |  |  |  |  |  |
| **N = 4,841** |  |  |  |  |  |
| Intercept | -0.24 [-0.95; 0.47] |  | 14.98 (6, 1010) | **< .001** | 0.79 |
| AUD criteria | -0.02 [-0.17; 0.14] |  | 0.05 (1, 64) | .825 | 0.98 |
| Weekend days versus weekdays<sup>#</sup> |  |  | 83.43 (1, 4834) | **< .001** | 1.79 |
| Weekend days | 0.58 [0.46; 0.71] |  |  |  |  |
| Weekdays | [Reference] |  |  |  |  |
| Cycle phases |  |  | 1.53 (4, 4834) | .192 |  |
| Menstrual | -0.01 [-0.21; 0.20] | .995 |  |  | 0.99 |
| Follicular | 0.20 [0.01; 0.40] | .044 |  |  | 1.22 |
| Ovulatory | 0.10 [-0.12; 0.31] | .393 |  |  | 1.10 |
| Mid-luteal | 0.11 [-0.07; 0.30] | .237 |  |  | 1.12 |
| Late luteal | [Reference] |  |  |  |  |
| **Progesterone/estradiol ratio** |  |  |  |  |  |
| **N = 72** |  |  |  |  |  |
| Intercept | 0.03 [-0.05; 0.12] |  | 0.10 (1, 64.45) | .759 |  |
| Time of blood collection | 4.27[-1.47; 10.02] |  | 2.21 (1, 64.56) | .142 |  |
| Cycle phases |  |  | 5.41 (4, 47.89) | **.001** |  |
| Menstrual | -0.06 [-0.11; -0.01] | **.009** |  |  |  |
| Follicular | -0.08 [-0.012; -0.04] | **< .001** |  |  |  |
| Ovulatory | -0.07 [-0.11; -0.03] | **.002** |  |  |  |
| Mid-luteal | -0.02 [-0.06; 0.17] | .262 |  |  |  |
| Late luteal | [Reference] |  |  |  |  |

AUD, alcohol use disorder. 95% CI, 95% confidence interval. p < .05 in bold

**Figure 1.**
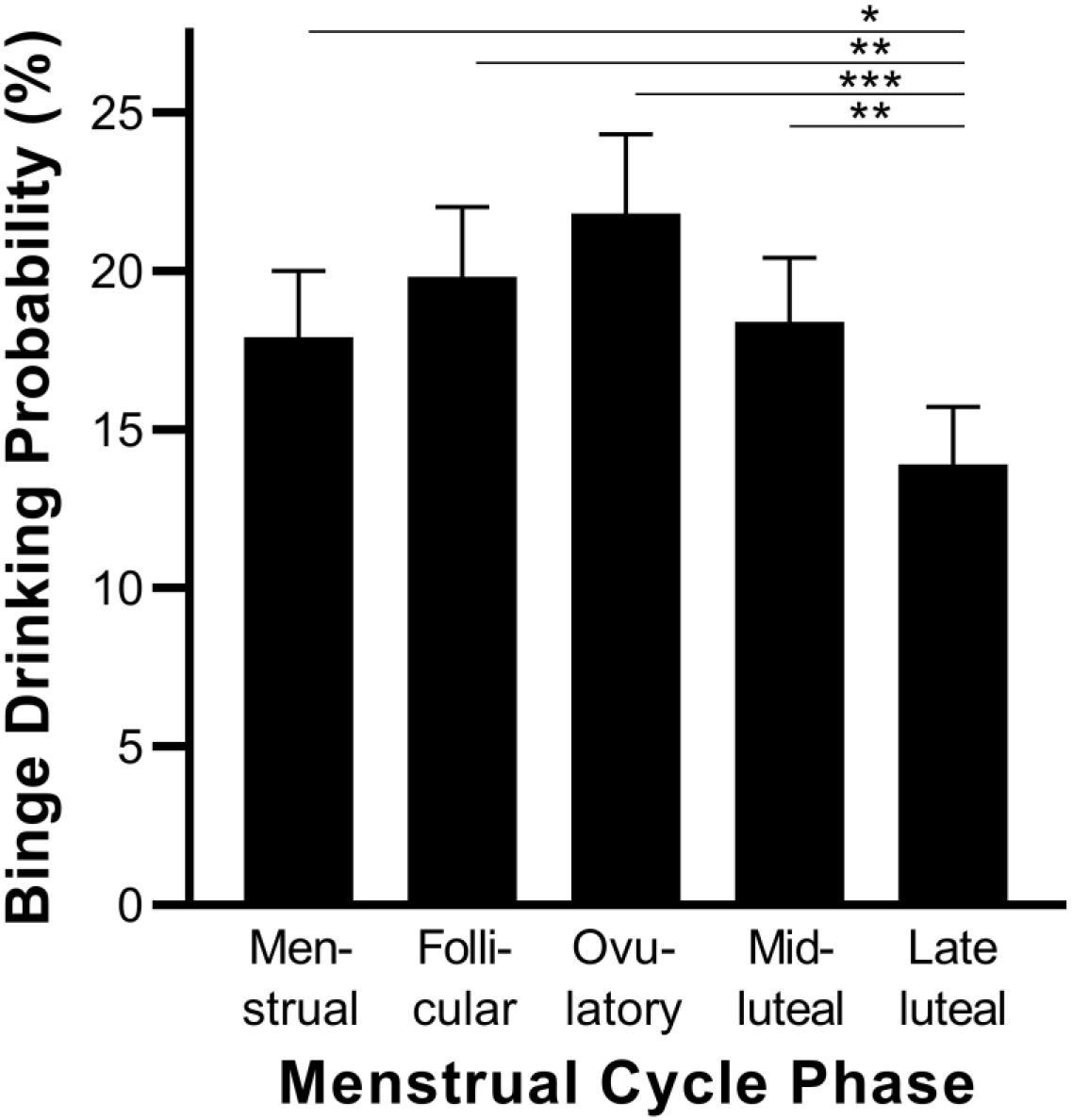
Binge Drinking Probability and Menstrual Cycle Phase In premenopausal naturally cycling females with alcohol use disorder, probability of binge drinking days varies across the menstrual cycle. N(cases) = 74; N(observations) per cycle phase: Menstrual 892, follicular 1,113, ovulatory 697, mid-luteal 1270, and late luteal 869. The graph shows marginal predicted probabilities and standard errors. *p < .05, **p < .01, ***p < .001.

### Males with AUD: Inverse Association between Progesterone/Estradiol Ratios and Alcohol Use

In males with AUD, higher progesterone/estradiol ratios were significantly associated with lower probabilities of binge drinking days and days with any alcohol use. An increase of 10 units in the progesterone/estradiol ratio was related to a decrease of 8% and 9% in the probability of binge drinking days and days with any alcohol use, respectively. The alcohol use patterns were significantly related to weekend days versus weekdays, and the binge drinking model was significantly affected by the number of positive AUD criteria (Table 3). The interactions of the progesterone/estradiol ratio with AUD criteria or weekend days versus weekdays did not reveal significant effects (data not shown).

**Table 3.** Multilevel Modeling Results in Males with AUD: Association of the Progesterone/Estradiol Ratios with Probability of Binge Drinking Days and Days with Any Alcohol Use Estimated in Separate Models

| | $\beta$ [95% CI] | F (df1, df2) | p | OR |
| --- | --- | --- | --- | --- |
| **Binge Drinking Day (yes/no)** |  |  |  |  |
| **N = 16,597** |  |  |  |  |
| Intercept | -1.78 [-2.46; -1.09] | 106.50 (4, 1544) | < .001 | 0.169 |
| AUD criteria | 0.12 [0.01; 0.23] | 4.32 (1, 228) | .039 | 1.126 |
| Weekend days versus weekdays |  | 418.94 (1, 16592) | < .001 | 2.276 |
| Weekend days | 0.82 [0.74; 0.90] |  |  |  |
| Weekdays | [Reference] |  |  |  |
| Time of blood collection | -0.00 [-0.04; 0.28] | 0.05 (1, 12892) | .832 | 0.997 |
| Progesterone/estradiol ratio | -0.01 [-0.02; -0.00] | 3.86 (1, 16592) | .049 | 0.992 |
| **Day with Any Alcohol Use (yes/no)** |  |  |  |  |
| **N = 16,597** |  |  |  |  |
| Intercept | 0.28 [-0.38; 0.95] | 93.99 (4, 1579) | < .001 | 1.329 |
| AUD criteria | 0.01 [-0.11; 0.12] | 0.02 (1, 229) | .903 | 1.007 |
| Weekend days versus weekdays |  | 371.76 (1, 16592) | < .001 | 2.079 |
| Weekend days | 0.73 [0.66; 0.81] |  |  |  |
| Weekdays | [Reference] |  |  |  |
| Time of blood collection | -0.00 [-0.04; 0.03] | 0.10 (1, 15648) | .756 | 0.995 |
| Progesterone/estradiol ratio | -0.01 [-0.02; -0.00] | 5.02 (1, 16592) | .025 | 0.991 |

95% CI, 95% confidence interval. p < .05 in bold

## Discussion

This study provides novel evidence for a role of progesterone relative to estradiol activities in problematic alcohol use of females and males with AUD. In line with our hypotheses, we established that in premenopausal naturally cycling females lower binge drinking probabilities and higher progesterone/estradiol ratios occur during the late luteal phase compared to the menstrual, follicular, and ovulatory phases. In males with AUD, we similarly identified an association of higher progesterone/estradiol ratios with lower probabilities of binge drinking and any alcohol use. These novel insights support the idea of pharmacologically increasing progesterone relative to estradiol activities for reducing problematic alcohol use in females and males with AUD.

Martel et al. (22) investigated 22 naturally cycling female university students (aged between 18 and 22 years, binge drinking on 4% of the recorded days) and found a lower likelihood of alcohol drinking in phases of high progesterone relative to estradiol saliva levels. Here, we successfully translated this finding to a sample of naturally cycling premenopausal females with a broader age range from 18 to 42 years, AUD, and a higher binge drinking probability of 21% of the recorded days. The finding suggests a protective effect of progesterone, which is in line with a previously reported correlation between higher progesterone serum levels and lower craving in female in-patients with AUD (10). However, contrary to our expectations based on the Martel et al. (22) study, the interaction between the menstrual cycle phases and weekend days versus weekdays was not significantly associated with alcohol use patterns. This finding may be due to a larger odds ratio for binge drinking during the weekend days versus weekdays of 10.34 in the Martel et al. (22) sample than in our sample with 2.18. The higher binge drinking probability and more work in the home office during the COVID-19 pandemic may lie at the bottom of the observed smaller differences between weekend days and weekdays (39).

Females with AUD often consume alcohol to cope with stress, depression, and anxiety (49) and the progesterone/estradiol ratio may be involved in the use of alcohol as a form of self-medication. For example, higher progesterone correlates with the lower cortisol stress response following a social stress test during the follicular phase (50), and in female rats, progesterone reduces anxiety-like behavior during nicotine withdrawal (51). The menstrual cycle status influences mood and emotion processing in addition to cortisol, adrenaline, and noradrenaline responses to stress (52-54). Studies in young social drinking females suggest that they drink alcohol to reduce negative affect during their perimenstrual and menstrual phases (55, 56). In accordance therewith, we observed an increase in binge drinking during the menses in our sample of relatively older premenopausal naturally cycling females with AUD. It should be noted that there are between-subject differences in sensitivity to varying cycle phases in naturally cycling females, and we likely recruited a cohort of females highly sensitive to hormones as AUD is associated with early abuse and stressful life events (57), impulsivity (58), and borderline personality features (59), all of which are linked to higher hormone sensitivity (60-64). Hence, the effects of progesterone on problematic alcohol use might depend on the individual drinking motives and the effects might be stronger in females with AUD than in social drinkers.

The changes in binge drinking across the menstrual cycle in females with AUD are supported by cycle-dependent alterations of brain reward sensitivity. The cycle phase influences brain reactivity to reward (19) with a stronger striatal activation during the follicular than during the luteal phase as demonstrated in a monetary reward paradigm (65) and in smoking-cue exposures (66). A study using a monetary incentive delay task revealed stronger responses to short-term rewards, greater activity in the dorsal striatum, and less activity in the right dorsolateral prefrontal cortex (indicative of less cognitive control of impulsivity) during the late follicular/ovulatory phase than during the mid-luteal phase and these effects were sensitive to estradiol (67). Accordingly, estrogen receptor 1 gene variations are associated with AUD (68, 69), and higher estradiol activities are related to an increase in alcohol use in adolescent and adult females (70, 71). Taken together, reward sensitivity is higher during the ovulatory phase (low progesterone/estradiol ratio) relative to the luteal phase (higher progesterone/estradiol ratio), which may underlie this study’s findings.

The observed changes in probability of binge drinking across the menstrual cycle may guide the development of treatments targeting the specific needs of females with AUD. Cycle phase-sensitive ecological momentary interventions or hormone therapy promise to yield significant advancements in the treatment of females with AUD. In terms of tobacco use disorder, smoking cessation is more often successful when started during the luteal (higher progesterone/estradiol ratio) than during the follicular phase (72-75). And initial evidence suggests an added benefit of naltrexone for drinking outcomes during cycle phases with high progesterone/estradiol ratios in premenopausal female heavy drinking smokers (76). As far as we know, such studies for AUD are lacking.

In males with AUD, we identified an association of higher progesterone/estradiol ratios with lower probabilities of binge drinking and any alcohol use. In line with this observation, a previous study found higher estradiol blood levels in males with AUD in comparison to controls (77). Moreover, a placebo-controlled study on cocaine-dependent individuals showed that the administration of progesterone reduces cue-induced craving and cortisol response. The effect sizes did not significantly differ between males and females, and safety measures indicated that progesterone was well-tolerated (16) (despite the lower progesterone blood concentrations in males versus females). Altogether, these findings highlight the potential role of progesterone as a treatment target for AUD also in males.

For decades, gestagen and estrogen modulators are used for various reasons (32), and preliminary study results indicate that progesterone treatment may be well-tolerated and safe in females and males with cocaine use disorder (16, 78). As identified in this study, the role of the progesterone/estradiol ratio in alcohol use might successfully lead to the development of novel treatment approaches for females and males with AUD targeting progesterone within the pregnenolone-progesterone-allopregnanolone pathway (79). Reduced progesterone levels are found in half of the females with AUD during the ovulation and luteal phase (80), and alcohol intoxication may increase progesterone levels in females (81). Pregnenolone and allopregnanolone modulate membrane and intracellular receptors including type-1 cannabinoid and γ-aminobutyric acid (GABA_A_) receptors. Both endocannabinoid and GABA signaling are involved in AUD (82, 83). Alcohol intoxication is related to an increase in allopregnanolone serum levels in female and male adolescents (81, 84), and ethanol injection increases brain allopregnanolone in male rats (85). By contrast, in a postmortem study, allopregnanolone and pregnenolone were strongly reduced in the cerebellum of males with AUD compared to controls (86).

### Strengths and Limitations

We investigated a large sample of females and males with mainly mild-to-moderate AUD across a broad age range. Self-reported alcohol use assessed retrospectively via the Timeline Followback Interview differs from daily reports (87, 88) with the latter providing ecologically more valid results through minimizing recall biases (25). Therefore, we employed advanced smartphone-based EMA technology across a 12-month period with sound compliance rates (75% answered queries on alcohol use) resulting in two large real-time intensively-sampled longitudinal data sets (21,438 smartphone entries) allowing hierarchical repeated measures analyses and ecological valid findings (89). Because of criticism related to self-report data on onset of menses (90), we used a standardized and established definition of menstrual cycle phases based on clear hormonal events (46) and verified the cycle phase classifications by quantifying serum levels of progesterone and estradiol. Future work needs to control for the effects of premenstrual dysphoric disorders, which is related to a paradoxical GABA_A_ response of negative affect following the exposure to allopregnanolone, especially during the late luteal phase (91). However, since we found the lowest probability of binge drinking during the late luteal phase, it is unlikely that our results were biased as a result of this GABA_A_-related factor.

### Conclusion and Outlook

We demonstrated a lower binge drinking probability in females during the late luteal phase (with higher progesterone/estradiol ratios) compared to the menstrual, follicular, and ovulatory cycle phases. Similarly, we established lower likelihoods of binge drinking and any alcohol use in males with higher progesterone/estradiol ratios. These findings indicate that the administration of progesterone to females and males with AUD may reduce problematic alcohol use. Moreover, the results suggest that menstrual cycle phase-dependent strategies can be beneficial in females with AUD. Future studies need to identify how drinking motives are involved in the here observed relationships and how brain (re)activities to alcohol cue exposure, incentive delay, and affective neural processing change across the menstrual cycle in females with AUD.

## Data Availability

All data produced in the present study are available upon reasonable request to the authors

## Role of Funding Source

The project was funded by the Deutsche Forschungsgemeinschaft (DFG, German Research Foundation) – Project-ID 402170461 – TRR265 (38). C.M. is a member of the research training group 2162 “Neurodevelopment and Vulnerability of the Central Nervous System” of the DFG (GRK2162 / 270949263). The funders had no role in the study design, data collection, analysis, decision to publish, or preparation of the manuscript.

## Acknowledgments

We thank the non-author collaborators of the ReCoDe Consortium: Felix Bermpohl, Christine Heim, Stefanie Kunas, Nina Romanczuk-Seiferth, Andreas Ströhle, Heiner Stuke (all Charité – Universitätsmedizin Berlin), Viktoria Arndt, Christian Beste, Hao Chen, Tanja Endraß, Sasha Frölich, Filippo Ghin, Stefan Kiebel, Clemens Kirschbaum, Michael Marxen, Wolfgang E. Nagel, Maximilian Pilhatsch, Sarah Schwöbel, Ann-Kathrin Stock (all Technische Universität Dresden), Gabriela Gan, Kristina Schwarz (all Central Institute of Mental Health Mannheim), and Anne Beck (Health and Medical University Potsdam).

## Author Contributions

Study concept and design: Hoffmann, Gerhardt, Mühle, Reinhard, D. Reichert, Bach, Sommer, Spanagel, Banaschewski, Deserno, Ebner-Priemer, Flor, Kirsch, Rietschel, Vollstädt-Klein, Walter, Meyer-Lindenberg, Rapp, Witt, Smolka, Heinz, Tost, Kiefer, M. Reichert, Lenz. Acquisition, analysis, or interpretation of data: All authors. Drafting of the manuscript: Hoffmann, Gerhardt, Lenz. Critical revision of the manuscript for important intellectual content: All authors. Statistical analysis: Hoffmann, Reinhard, D. Reichert, M. Reichert. Obtained funding: Sommer, Spanagel, Banaschewski, Deserno, Ebner-Priemer, Flor, Kirsch, Rietschel, Vollstädt-Klein, Walter, Meyer-Lindenberg, Rapp, Smolka, Heinz, Tost, Kiefer, Lenz. Administrative, technical, and/or material support: Mühle, Kornhuber, Banaschewski, Deserno, Ebner-Priemer, Flor, Kirsch, Rietschel, Vollstädt-Klein, Walter, Meyer-Lindenberg, Rapp, Smolka, Heinz, Tost, Kiefer, Lenz. Study supervision: Banaschewski, Deserno, Ebner-Priemer, Flor, Kirsch, Rietschel, Vollstädt-Klein, Walter, Rapp, Smolka, Heinz, Tost, Kiefer, Lenz.

## Disclosures

Ulrich W. Ebner-Priemer is a consultant for Boehringer Ingelheim.

## Footnotes

1 As we refer to sex rather than gender, we chose to use the terms female and male instead of the gender-related terms woman and man.

## Notes

### Author Declarations

The project was approved by the local ethics committees of Charite - Universitaetsmedizin Berlin, Technical University Dresden, and Heidelberg University.

