## Supplementary Information for "Progesterone/Estradiol Ratio Is Related to Real-Life Alcohol Consumption in Alcohol Use Disorder in a Sex- and Menstrual Cycle Phase-Dependent Manner"

**Supplementary Figure S1.** Study Design

**Supplementary Figure S2.** Progesterone/Estradiol Ratio and Menstrual Cycle Phase

**Supplementary Figure S3.** % of Binge Drinking Days Across the Menstrual Cycle

**Supplementary Table S1.** Definition of Menstrual Cycle Phases According to Cycle Length in Days

**Supplementary Table S2.** Multilevel Modeling Results in Premenopausal Naturally Cycling Females with AUD: Association of Menstrual Cycle Day with Probability of Binge Drinking Days and Days with Any Alcohol Use Estimated in Separate Models

**Supplementary Figure S1.** Study Design

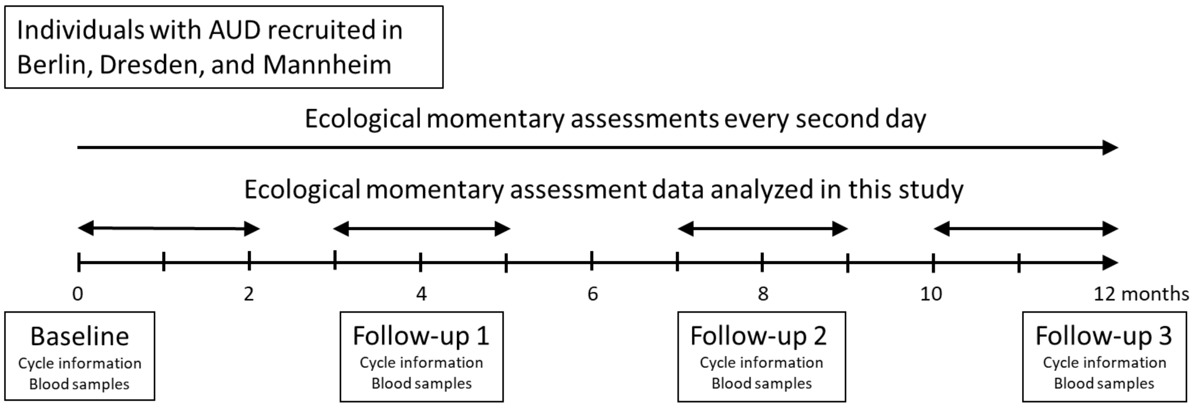


Caption Supplementary Figure S1. The chart illustrates how the study design combines ecological momentary assessments (EMA) with blood samples. The females and males with alcohol use disorder (AUD) were followed up to 12 months with EMA every second day and attended onset assessments at baseline, follow-up 1 after 4 months, follow-up 2 after 8 months, and follow-up 3 after 12 months. For this study, we analyzed EMA data from several periods: 56 days (≙ 2 menstrual cycles) after baseline, 28 days before and after follow-ups 1 and 2, and 56 days before follow-up 3.

**Supplementary Figure S2.** Progesterone/Estradiol Ratio and Menstrual Cycle Phase

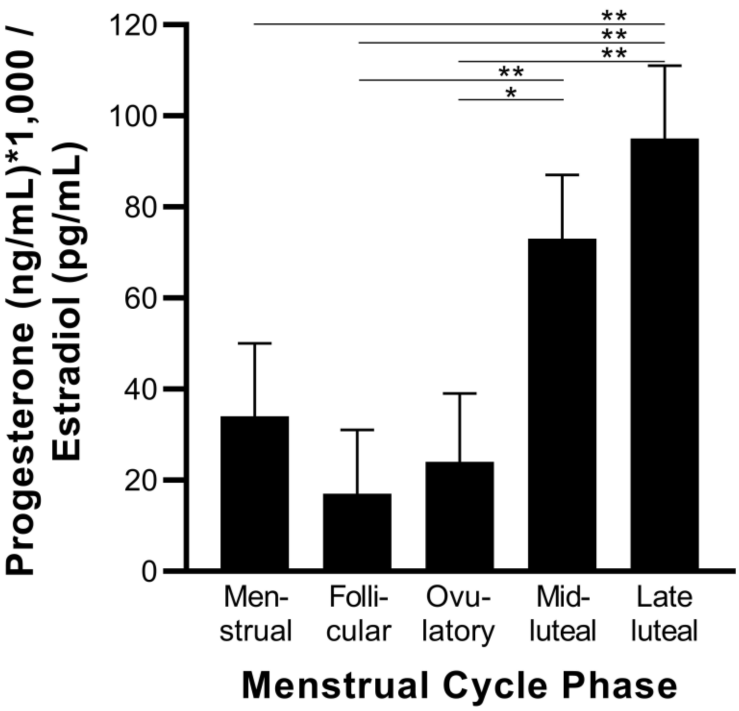


Caption Supplementary Figure S2. In premenopausal naturally cycling females with alcohol use disorder, the mean progesterone/estradiol ratio varies across the menstrual cycle. N(cases) / N(observations) per cycle phase: Menstrual 12 / 13, follicular 15 / 17, ovulatory 13 / 13, mid-luteal 15 / 17, and late luteal 12 / 12. The graph shows estimated marginal means and standard errors. *p < .05, **p < .01.

**Supplementary Figure S3.** % of Binge Drinking Days Across the Menstrual Cycle

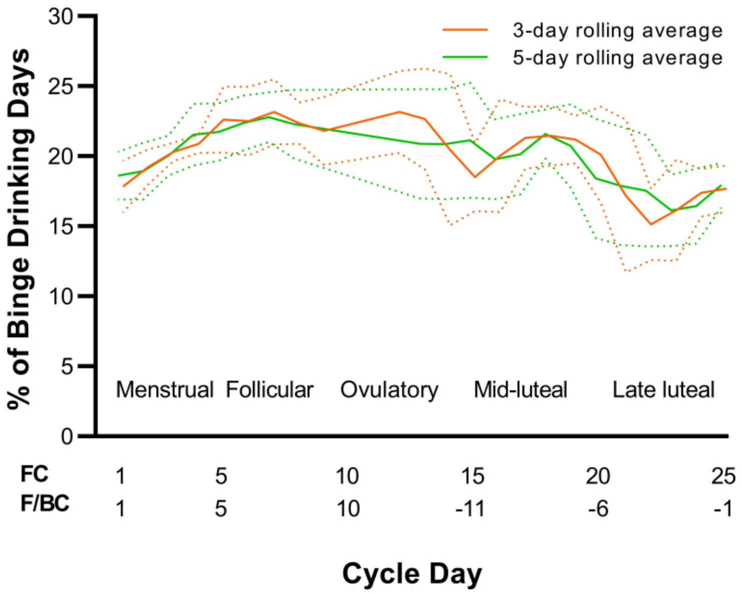


Caption Supplementary Figure S3. The graph shows the raw observed probabilities of binge drinking per menstrual cycle day and the standard deviations due to the 3-day and 5-day rolling average transformation, which we chose for visualization of the cyclic changes. Methods to determine cycle day (1): FC, forward count, F/BC forward/backward count.

**Supplementary Table S1.** Definition of Menstrual Cycle Phases According to Cycle Length in Days

| Total cycle length (days) | Days of cycle phases | | | | |
|---|---|---|---|---|---|
| | Menstrual | Follicular | Ovulatory | Mid-luteal | Late luteal |
| 22 | 1-5 | 6 | 7-10 | 11-17 | 18-22 |
| 25 | 1-5 | 6-9 | 10-13 | 14-20 | 21-25 |
| 28 | 1-5 | 6-12 | 13-16 | 17-23 | 24-28 |
| 31 | 1-5 | 6-15 | 16-19 | 20-26 | 27-31 |
| 34 | 1-5 | 6-18 | 19-22 | 23-29 | 30-34 |

Method to determine menstrual cycle phase according to the individual cycle length.

**Supplementary Table S2.** Multilevel Modeling Results in Premenopausal Naturally Cycling Females with AUD: Association of Menstrual Cycle Day with Probability of Binge Drinking Days and Days with Any Alcohol Use Estimated in Separate Models

| | F (df1, df2) | p |
|---|---|---|
| **Binge Drinking Day (yes/no)** | | |
| **N = 4,841** | | |
| Intercept | 25.30 (4, 521) | **< .001** |
| AUD criteria | 1.52 (1, 66) | .222 |
| Weekend days versus weekdays | 96.00 (1, 4359) | **< .001** |
| Cycle day | 1.29 (1, 4359) | .257 |
| Squared cycle day | 4.17 (1, 4359) | **.041** |
| **Day with Any Alcohol Use (yes/no)** | | |
| **N = 4,841** | | |
| Intercept | 19.17 (4, 499) | **< .001** |
| AUD criteria | 0.07 (1, 63) | .789 |
| Weekend days versus weekdays | 75.43 (1, 4359) | **< .001** |
| Cycle day | 0.00 (1, 4359) | .966 |
| Squared cycle day | 0.60 (1, 4359) | .438 |

$p < .05$ in bold
